# Tracing DAY-ZERO and Forecasting the COVID-19 Outbreak in Lombardy, Italy: A Compartmental Modelling and Numerical Optimization Approach

**DOI:** 10.1101/2020.03.17.20037689

**Authors:** Lucia Russo, Cleo Anastassopoulou, Athanasios Tsakris, Gennaro Nicola Bifulco, Emilio Fortunato Campana, Gerardo Toraldo, Constantinos Siettos

## Abstract

Italy became the second epicenter of the novel coronavirus disease 2019 (COVID-19) pandemic after China, surpassing by far China’s death toll. The disease swept through Lombardy, which remained in lockdown for about two months, starting from the 8th of March. As of that day, the isolation measures taken in Lombardy were extended to the entire country. Here, assuming that effectively there was one case “zero” that introduced the virus to the region, we provide estimates for: (a) the DAY-ZERO of the outbreak in Lombardy, Italy; (b) the actual number of asymptomatic infected cases in the total population until March 8; (c) the basic reproduction number (*R*_0_) based on the estimation of the actual number of infected cases. To demonstrate the efficiency of the model and approach, we also provide a tentative forecast two months ahead of time, i.e. until May 4, the date on which relaxation of the measures commenced, on the basis of the COVID-19 Community Mobility Reports released by Google on March 29.

**Methods:** To deal with the uncertainty in the number of actual asymptomatic infected cases in the total population, we address a modified compartmental Susceptible/ Exposed/ Infectious Asymptomatic/ Infected Symptomatic/ Recovered/ Dead (SEIIRD) model with two compartments of infectious persons: one modelling the cases in the population that are asymptomatic or experience very mild symptoms and another modelling the infected cases with mild to severe symptoms. The parameters of the model corresponding to the recovery period, the time from the onset of symptoms to death and the time from exposure to the time that an individual starts to be infectious, have been set as reported from clinical studies on COVID-19. For the estimation of the DAY-ZERO of the outbreak in Lombardy, as well as of the “effective” per-day transmission rate for which no clinical data are available, we have used the proposed SEIIRD simulator to fit the numbers of new daily cases from February 21 to the 8th of March. This was accomplished by solving a mixed-integer optimization problem. Based on the computed parameters, we also provide an estimation of the basic reproduction number *R*_0_. To examine the efficiency of the model and approach, we ran the simulator to “forecast” the epidemic two months ahead of time, i.e. from March 8 to May 4. For this purpose, we considered the reduction in mobility in Lombardy as released on March 29 by Google COVID-19 Community Mobility Reports, and the effects of social distancing and of the draconian measures taken by the government on March 20 and March 21, 2020.

**Results:** Based on the proposed methodological procedure, we estimated that the expected DAY-ZERO was January 14 (min-max rage: January 5 to January 23, interquartile range: January 11 to January 18). The actual cumulative number of asymptomatic infected cases in the total population in Lombardy on March 8 was of the order of 15 times the confirmed cumulative number of infected cases, while the expected value of the basic reproduction number *R*_0_ was found to be 4.53 (min-max range: 4.40-4.65).

The model approximated adequately two months ahead of time the evolution of reported cases until May 4, the day on which the phase I of the relaxation of measures was implemented over all of Italy.

## Introduction

The butterfly effect in chaos theory underscores the sensitive dependence on initial conditions, highlighting the importance of even a small change in the initial state of a nonlinear system. The emergence of a novel coronavirus, SARS-CoV-2, that caused a viral pneumonia outbreak in Wuhan, Hubei province, China in early December 2019 has evolved into the COVID-19 acute respiratory disease pandemic due to its alarming levels of spread and severity, with more than 3.5 million cases and 250,000 deaths globally, as of May 7, 2020 ([1, 2]). The seemingly far from the epicenter, old continent became the second-most impacted region after Asia Pacific, mostly as a result of a dramatic divergence of the epidemic trajectory in Italy first, where there have been 214,457 total confirmed infected cases and 29,684 deaths, and then in Spain where there have been 220,325 total confirmed infected cases and 25,857 deaths, as of May 7, 2020 ([1, 2]).

The second largest outbreak outside of mainland China officially started on January 31, 2020, after two Chinese visitors staying at a central hotel in Rome tested positive for SARS-CoV-2; the couple remained in isolation and was declared recovered on February 26 [3]. A 38-year-old man repatriated back to Italy from Wuhan who was admitted to the hospital in Codogno, Lombardy on February 21 was the first secondary infection case (“patient 1”). “Patient 0” was never identified by tracing the first Italian citizen’s movements and contacts. In less than a week, the explosive increase in the number of cases in several bordering regions and autonomous provinces of northern Italy placed enormous strain on the decentralized health system. Following a dramatic spike in deaths from COVID-19, Italy transformed into a “red zone”, and the movement restrictions were expanded to the entire country on the 8th of March. All public gatherings were cancelled and school and university closures were extended through at least the next month.

In an attempt to assess the dynamics of the outbreak for forecasting purposes, it is important to estimate epidemiological parameters that cannot be computed directly based on clinical data, such as the transmission rate of the disease and the basic reproduction number, *R*_0_, defined as the expected number of exposed cases generated by one infected case in a population where all individuals are susceptible, many mathematical modelling studies have already appeared since the first confirmed COVID-19 case. The first models mainly focused on the estimation of the basic reproduction number *R*_0_ using dynamic mechanistic mathematical models ([4, 5, 6, 7]), but also simple exponential growth models (see e.g. [8, 9]). Compartmental epidemiological models like SIR, SIRD, SEIR and SEIRD have been proposed to estimate other important epidemiological parameters, such as the transmission rate and for forecasting purposes (see e.g. [7, 10]). Other studies have used metapopulation models, which include data of human mobility between cities and/or regions to forecast the evolution of the outbreak in other regions/countries far from the original epicenter in China [4, 11, 12, 6], including the modelling of the influence of travel restrictions and other control measures in reducing the spread ([13].

Among the perplexing problems that mathematical models face when they are used to estimate epidemiological parameters and to forecast the evolution of the outbreak, two stand out: (a) the uncertainty regarding the DAY-ZERO of the outbreak, the knowledge of which is crucial to assess the stage and dynamics of the epidemic, especially during the first growth period, and (b) the uncertainty that characterizes the actual number of asymptomatic infected cases in the total population (see e.g. [14]).

At this point we should note that what is done until now with dynamical epidemiological models is the investigation of several scenarios including different “DAYS-ZERO” or just fixing the DAY-ZERO and run different levels of asymptomatic cases e.t.c. To cope with the above problems, we herein address a methodological framework that provides estimates for the DAY-ZERO of the outbreak and the number of the asymptomatic cases in the total population in a systematic way. Towards this goal, and for our demonstrations, we address a conceptually simple SEIRD model with two compartments, one modelling the asymptomatic infected cases in the population and another modelling the part of the infected cases that will experience mild to severe symptoms, a significant share of which will be hospitalized, admitted to intensive care units (ICUs) or die from the disease. The proposed approach is applied to Lombardy, the epicenter of the outbreak in Italy. Furthermore, we provide a two-months ahead of time forecast from March 8 (the day of lockdown of all Italy) to May 4 (the first day of the relaxation of the strict isolation measures). The above tasks were accomplished by the numerical solution of a mixed-integer optimization problem using the publicly available data of daily new cases for the period February 21- March 8, and the COVID-19 Community Mobility Reports released by Google on March 29.

## Methodology

### The modelling approach

We address a compartmental SEIIRD model that includes two categories of infected cases, namely the asymptomatic (unknown) cases in the total population and the cases that develop mild to more severe symptoms, a significant share of which are hospitalized, admitted to ICUs and a part of them dies. In agreement with other studies and observations, our modelling hypothesis is that the confirmed cases of infected are only a (small) subset of the actual number of asymptomatic infected cases in the total population [6, 14, 7]. Regarding the confirmed cases of infected as of February 21, a study conducted by the Chinese CDC which was based on a total of 72,314 cases in China, found that about 80.9% of the cases were mild and could recover at home, 13.8% severe and 4.7% critical [15].

On the basis of the above findings, in our modelling approach, it is assumed that the asymptomatic or very mildly symptomatic cases recover from the disease relatively soon and without medical care, while for the other category of infected, on average their recovery lasts longer than the non-confirmed, they may also be hospitalized, admitted to ICUs or die from the disease.

Based on the above, let us consider a well-mixed population of size *N*. The state of the system at time t, is described by (see also Figure 1 for a schematic) *S*(*t*) representing the number of susceptible persons, *E*(*t*) the number of exposed, *I*(*t*) the number of asymptomatic infected persons in the total population who experience very mild or no symptoms and recover relatively soon without any other complications, *I_c_*(*t*) the number of infected cases who may develop mild to more severe symptoms and a significant part of them is hospitalized, admitted to ICUs or dies, *R*(*t*) the number of asymptomatic cases in the total population that recover, *R_c_(t*) the number of the recovered cases that come from the compartment of *I_c_* and *D*(*t*) the reported number of deaths. For our analysis, and for such a short period, we assume that the total number of the population remains constant. Based on demographic data, the total population of Lombardy is *N* = 10m; its surface area is 23,863.09 *kmq* and the population density is ~422 (Inhabitants/Kmq).

The rate at which a susceptible (*S*) becomes exposed (*E*) to the virus is proportional to the density of infectious persons *I*. The proportionality constant is the “effective” disease transmission rate, say 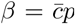, where 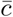 is the average number of contacts per day and p is the probability of infection upon a contact between a susceptible and an infected. Our main assumption here is that only a fraction, say *∊* of the actual number of exposed cases *E* will experience mild to more severe symptoms denoted as *I_c_*(*t*) and a significant part of them will be hospitalized, admitted to ICUs or die. Thus, we assume that the infected persons that belong to the compartment *I_c_* go into quarantine at home or they are hospitalized, and, thus, it is assumed that for any practical means they don’t transmit further the disease. Here, it should be noted that a wide testing policy may also result in the identification of asymptomatic cases belonging to the compartment *I* that would then be assigned to compartment *I_c_*. However, as a generally reported rule in Italy, tests were conducted only for those who presented for treatment with symptoms like fever and coughing. Thus, people who did not seek medical attention were tested very scarcely [16, 17, 18]. Thus, for any practical means the compartment *I_c_* reflects the reported confirmed infected cases.

**Figure 1:**
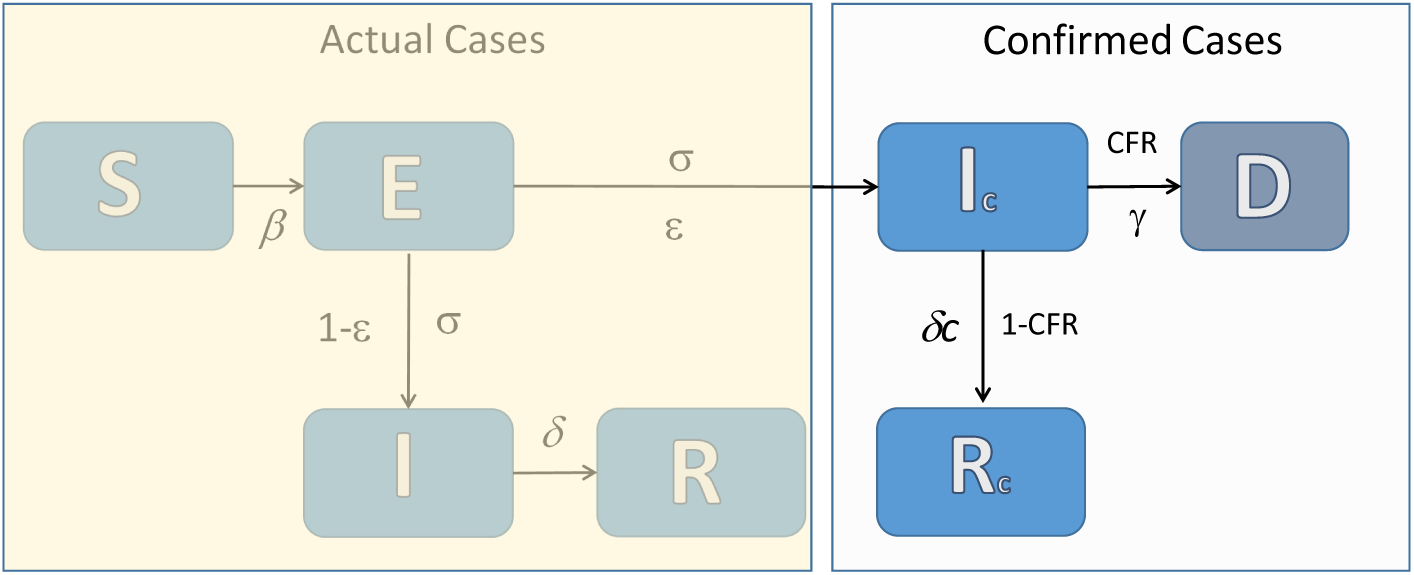
A schematic of the proposed compartmental SEIIRD model. The actual number of cases is unknown.

A fraction of the *I_c_* cases that is given by the fatality ratio 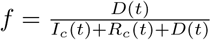 dies with a mortality rate *γ* the inverse of which is the average time from the onset of symptoms to death, while the remaining part ((1 − *f*)) of the *I_c_* compartment recovers with a rate *δ_c_*, the inverse of which corresponds to the average time from the onset of symptoms to full recovery.

We note that while more compartments could conceptually be included, we aimed at keeping a low level of complexity in order to avoid the introduction of more parameters, and thus a model that would suffer from the “curse of dimensionality”.

At this point we should note that on March 8, the date of the general lockdown, the number of confirmed infected cases was 3,372, the number of cases in ICUs was 399 and the number of hospitalized persons was 2,616 [19]. That is, until March 8, the number of confirmed cases was approximately equal to the number of hospitalized cases and the cases that were admitted to ICUs. Therefore, until March 8, any difference between the asymptomatic cases as represented by our model by the compartment *I* and the compartment *I_c_* would approximately reflect a level of under-reporting of the actual asymptomatic cases in the total population. We should also note that in the available data of reported cases [19] there is no distinction between cases that recovered at home and those that recovered at and were dismissed from hospitals. Thus, in the absence of such information, if one were to consider as a separate category the cases that are hospitalized, an extra parameter would have to be introduced (the fraction of recovered cases dismissed from hospitals). On one hand, such a piece of information is not available, and, on the other, such an attempt would add an extra degree of freedom that would need calibration or to be fixed at a certain value; however, due to the small size of the data and the “curse of dimensionality”, this would also introduce unnecessary computational burden and further modelling uncertainty.

Thus, our discrete mean field compartmental SEIIRD model reads:

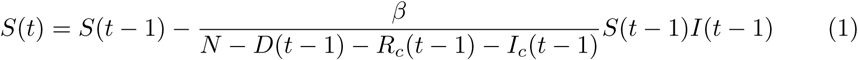

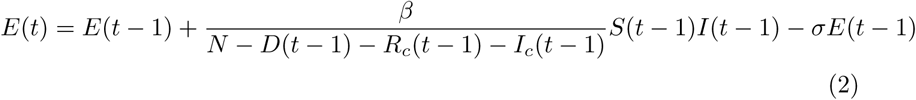

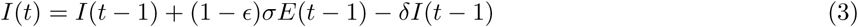

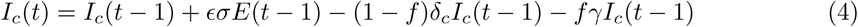

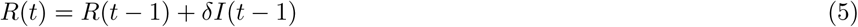

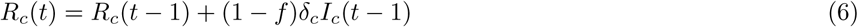

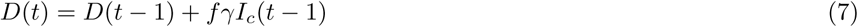

The above system is defined in discrete time points *t* = 1, 2,…, with the corresponding initial condition at the very start of the outbreak (DAY-ZERO): *S*(0) = *N −* 1, *I*(0) = 1, *E*(0) = 0 *I_c_*(0) = 0, *R*(0) = 0, *R_c_*(0) = 0, *D*(0) = 0.

The parameters of the model are:

- *β*(*d*^−1^) is the “effective” transmission rate of the disease,
- *σ*(*d^-1^*) is the average per-day “effective” rate at which an exposed person becomes infectious,
- *δ*(*d*^−1^) is the average per-day “effective” recovery rate within the group of asymptomatic cases in the total population,
- *δ_c_*(*d*^−1^) is the average per-day “effective” recovery rate within the subset of the *I_c_* infected cases that finally recover,
- *γ*(*d*^−1^) is the average per-day “effective” mortality rate within the subset of *I_c_* infected cases that finally die,
- *f* is the probability that a *I_c_* case will die. Here, this, is given by the “emergent” case fatality ratio, computed as 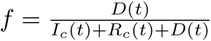,
- *∊*(*d*^−1^) is the fraction of the actual (all) cases of exposed in the total population that enter to the compartment *I_c_*.

Here, we should note the following: as new cases of recovered and dead at each time *t* appear with a time delay (which is generally unknown but an estimate can be obtained by clinical studies) with respect to the corresponding infected cases, the above per-day rates are not the actual ones; thus, they are denoted as “effective/apparent” rates.

The values of the epidemiological parameters *σ*, *δ, δ_c_, γ* that were fixed in the proposed model were chosen based on clinical studies.

In particular, in many studies that use SEIRD models, the parameter *σ* is set equal to the inverse of the mean incubation period (time from exposure to the development of symptoms) of a virus. *However, the incubation period does not generally coincide with the time from exposure to the time that someone starts to be infectious*. Regarding COVID-19, it has been suggested that an exposed person can be infectious well before the development of symptoms [20]. With respect to the incubation period for SARS-CoV-2, a study in China [21] suggests that it may range from 2-14 days, with a median of 5.2 days. Another study in China, using data from 1,099 patients with laboratory-confirmed 2019-nCoV ARD from 552 hospitals in 31 provinces/provincial municipalities suggested that the median incubation period is 4 days (interquartile range, 2 to 7). In our model, as explained above, 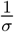 represents the period from exposure to the onset of the contagious period. Thus, based on the above clinical studies, for our simulations, we have set 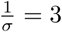.

Regarding the recovery period, in a study that is based on 55,924 laboratory-confirmed cases, the WHO-China Joint Mission has reported a median time of 2 weeks from onset to clinical recovery for mild cases, and 3-6 weeks for severe or critical cases [22]. Based on the above, and on the fact that within the subset of confirmed cases the mild cases are the 81% [15], we have set the recovery period for the confirmed cases’ compartment to be *δ_c_* = 1/21 in order to balance the recovery period with the corresponding characterization of the cases (mild, severe/critical). The average recovery period of the unreported/non-confirmed part of the infected population, which in our assumptions experiences the disease like the flu or a common cold, is set equal to one week [23], i.e. we have set *δ* = 1*/*7. This choice is based also on reports on the serial interval of COVID-19. The serial interval of COVID-19 is defined as the time duration between a primary case-patient (infector) having symptoms and a secondary case-patient having again symptoms. For example, it has been reported that the serial interval for COVID is estimated at 4.4–7.5 days [24]; for the case of Lombardy, the average serial number has been estimated to be 6.6 days [25]. In our model, the 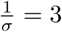 period refers to the period from exposure to the onset of the contagiousness. In this period, obviously there are no symptoms. Thus, the serial interval in our model is 7 days (this is the average number of days in which an infectious becomes recovered and no longer transmits the disease). Importantly, there are studies (see e.g. Nushiura et al. [26]) suggesting that a substantial proportion of secondary transmission may occur prior to illness onset. Thus, the 7 days period that we have taken as the average period that an infectious person can transmit the disease before he/she recovers, reflects exactly this period; it refers to the serial interval for the cases that are asymptomatic and for cases with mild symptoms.

Finally, the median time from the onset of symptoms until death for Italy has been reported to be eight days [27], thus in our model we have set *γ* = 1/8.

We have set *f* = 11% for the optimization. For the forecasting (i.e. for the period March 9 to May 4), the value of *f* was not fixed but it was computed dynamically each day *t* through the model simulations as 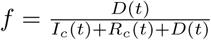.

The transmission rate *β*, as it cannot be obtained in general by clinical studies, but only by mathematical models, was estimated through the optimization process.

Regarding DAY-ZERO in Lombardy, that is also unknown and estimated by the optimization process, what has been officially reported is just the date on which the first infected person was confirmed to be positive for SARS-CoV-2. That day was February 21, 2020, which is the starting date of public data release of confirmed cases.

### Estimation of the DAY-ZERO of the outbreak, the scale of data uncertainty and the disease transmission

The DAY-ZERO of the outbreak, the per-day “effective” transmission rate *β*, and the ratio *∊* were computed by the numerical solution of a mixed-integer optimization problem with the aid of genetic algorithms to fit the reported data of daily new cases (see the discussion in [28]) from February 21 to March 8, the day of the lockdown of Lombardy.

As already mentioned, on March 8, the number of confirmed infected cases in the population was 3,372, the number of cases in ICUs was 399 and the number of hospitalized persons was 2,616 [19]. That is, until March 8 the number of mild and severe cases that were hospitalized and admitted to ICUs was approximately equal to the number of confirmed infected cases. Thus, for the period of calibration, it is reasonable to assume that for any practical means the number of confirmed cases was approximately the same as for those that experienced mild to more severe symptoms and were admitted for medical care. Thus, until March 8, the parameter *∊* reflects also the level of under-reporting of the asymptomatic cases in the total population.

Here, for our computations, we have used the genetic algorithm “ga” provided by the Global Optimization Toolbox of Matlab [5] to minimize the following objective function:

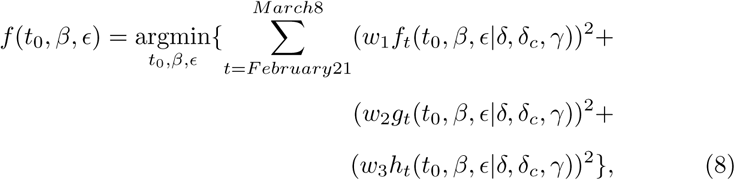

where,

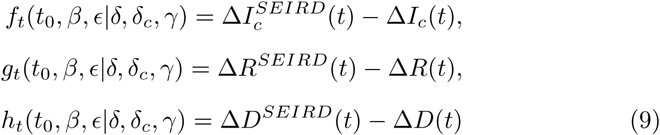

∆*x^seird^*(*t*), (*X* = *I*, *R*, *D*) are the new cases resulting from the SEIRD simulator at time *t*.

The weights *w*_1_, *w*_2_, *w*_3_ correspond to scalars serving in the general case as weights to the relevant functions for balancing the different scales between the number of infected, recovered cases and deaths.

At this point we should note that the above optimization problem may in principle have more than one nearby optimal solutions, which may be attributed to the fact that the tuning of both DAY-ZERO and the transmission rate may in essence result in nearby values of the objective function. In order to quantify the above uncertainty in the optimization procedure, we created a grid of initial guesses within the intervals in which the optimal estimates were sought: for the DAY-ZERO (*t*_0_) we used a step of days within the interval December 27, 2019 until the 5th of February, 2020 i.e. ±20 days around the 16th of January, for *β* we used a step of 0.05 within the interval (0.3, 0.9) and for *∊* we used a step of 0.02 within the interval (0.01, 0.29). The numerical optimization procedure was repeated 48 times for each combination of initial guesses. For our computations, we kept the best fitting outcome for each combination of initial guesses. Next, in order to reveal structured patterns of distributions vs. uniformly random distributions, we fitted the resulting probability distributions of the optimal values using several functions, including the Normal, Log-normal, Weibull, Beta, Gamma, Burr, Exponential and Birnbaum-Saunders distributions and kept the one resulting in the maximum Log-likelihood (see in the Supporting Information for more details). For the computed parameters of the corresponding best distributions, we also provide the corresponding 95% confidence intervals. Such fitting can demonstrate that the obtained values are not uniformly distributed.

Finally, we have run simulations based on all obtained values to assess the efficiency of the model and obtained results and the forecasting uncertainty until May 4.

For our computations, we used the parallel computing toolbox of Matlab 2020a [29] utilizing 6 INTEL XEON CPU X5650 cores at 2.66GHz.

### Estimation of the basic reproduction number R_0_ from the SEIIRD model

Here, we note that we provide an estimation of the basic reproduction number *R*_0_ based on the estimation of the total number of (asymptomatic) infected cases in the population. Thus, it is expected that the estimated *R*_0_ will be larger than the ones reported using just the confirmed number of cases; the latter may underestimate the actual *R*0.

Initially, when the spread of the epidemic starts, all the population is considered to be susceptible, i.e. *S* ≈ *N*. On the basis of this assumption, we computed the basic reproduction number based on the estimates of the epidemiological parameters computed using the data from the 21st of February to the 8th of March with the aid of the SEIIRD model given by Eq.(1)-(7) as follows.

Note that there are three infected compartments, namely *E, I, I_c_* and two of them (*E*,*I*) determine the outbreak. Thus, considering the corresponding equations given by Eq.(2),(3),(4), and that at the very first days of the epidemic *S* ≈ *N* and *D* ≈ 0, the Jacobian of the system as evaluated at the disease-free state reads:

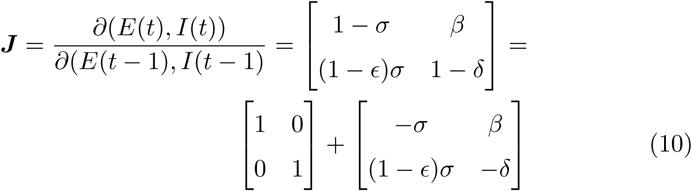

The eigenvalues (that is the roots of the characteristic polynomial of the Jacobian matrix) dictate if the disease-free equilibrium is stable or not, that is if an emerging infectious disease can spread in the population. In particular, the disease-free state is stable, meaning that an infectious disease will not result in an outbreak, if and only if all the norms of the eigenvalues of the Jacobian ***J*** of the discrete time system are bounded by one. Jury's stability criterion [30] (the analogue of Routh-Hurwitz criterion for discrete-time systems) can be used to determine the stability of the linearized discrete time system by analysis of the coefficients of its characteristic polynomial. The characteristic polynomial of the Jacobian matrix reads:

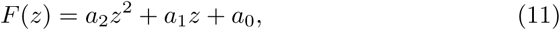

where

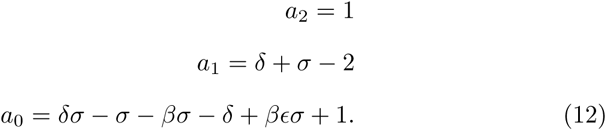

The necessary conditions for stability read:

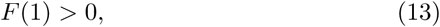

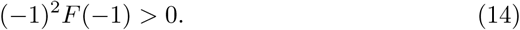

The sufficient conditions for stability are given by the following two inequalities:

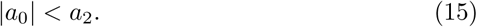

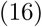

The first inequality (13) results in the necessary condition

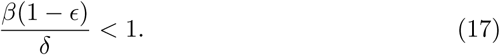

It can be shown that the second necessary condition (14) and the sufficient condition (15) are always satisfied for the range of values of the epidemiological parameters considered here.

Thus, the necessary condition (17) is also a sufficient condition for stability. Hence, the disease-free state is stable, if and only if, condition (17) is satisfied.

Note that in this necessary and sufficient condition (17), the fraction 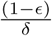 is the average infection time of the compartment I. Thus, the above expression reflects the basic reproduction number R0 which is qualitatively defined by 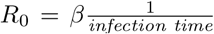. Hence, our model results in the following expression for the basic reproduction number:

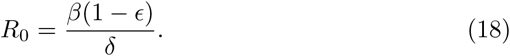

Note that for *∊* = 0, the above expression simplifies to *R*_0_ for the simple SIR model.

### Forecasting

As discussed, we used the proposed approach to forecast the evolution of the pandemic in Lombardy from March 8 to May 4, i.e. from the first day of lockdown to the first day of the relaxation of the social isolation.

Our estimation regarding the as of March 8 reduction of the “effective” transmission rate was based on the combined effects of prevention efforts and behavioral changes. In particular, our estimation was based on (a) the COVID-19 Community Mobility Reports released by Google on March 29 [31], and, (b) an assessment of the synergistic effects of such control measures as the implementation of preventive containment in workplaces, stringent “social distancing”, and the ban on social gatherings, as well as the public awareness campaign prompting people to adopt cautious behaviors to reduce the risk of disease transmission (see also [32, 33, 34, 35]). The effect of the distribution of contacts at home, work, when travelling, and during leisure activities can be also assessed. For example, based on an analysis for the social contacts and mixing patterns relevant to the spread of infectious diseases that was conducted in various countries, it has been found that for Italy, around 20-23% of all physical social contacts during a day are attributed to workplaces, around 17-18% to schools, 2-3% to transportation, 20% to leisure activities, 15-18% to home, 15% to other activities (contacts made at locations other than home, work, school, travel, or leisure) and a 7-8% to contacts made at multiple other locations during the day, not just at a single location [36].

On the basis of the Google COVID-19 Community Mobility Report released on March 29 [31], the average reduction in the mobility in Lombardy during the period February 16-March 8, compared to the period before February 16, was ~15% in retail & recreation activities, ~20% in transit stations, ~12% in workplaces, while it was increased in parks by ~ 11% and was almost the same in groceries and pharmacies. In the period March 9 to March 20, the mobility was reduced by an average of ~73% in retail & recreation activities, by ~75% in transit stations, by ~55% in workplaces, by ~58% in parks and by ~ 32% in groceries and pharmacies. Thus, taking into account the coarse effect of different activities in the physical contact [36], the average reduction was of the order of ~40% in mobility when compared to the period February 21-March 8. In fact, on March 17, based on the release of mobile phone data, the vice-president of Lombardy, announced that the average mobility in the region (for distances more than 500 meters) had been reduced by a ~50% with respect to the period before February 20 [37]. On March 20, the government announced the implementation of even stricter measures that included the closure of all public and private offices, closing all parks, walking only around the residency and not even in pairs, and the prohibition of mobility to second houses [38, 39]. According to the Google COVID-19 Community Mobility Reports [31], from March 20-21 until April 30, activities were reduced by an average of ~87% in retail & recreation activities, by ~84% in transit stations, by ~70% in workplaces, by ~80% in Parks and by ~ 50% in groceries and pharmacies. Thus, taking into account the coarse effect of different activities in the physical contact [36], the average reduction was of the order of ~65% in the mobility when compared to the period of February 21-March 8.

A further reduction may be attributed to behavioral changes. For example, it has been shown that social distancing and cautiousness reduce the disease transmission rate by about 20% [34]. Thus, based on the above, it is reasonable to consider a (1-0.4)(1-0.2) (an average of 40% contribution of the reduction of the mobility and a 20% for the effect of social distancing) reduction in the effective transmission rate for the period March 8-March 19 as compared with the period February 21-March 8. For the period of March 20-21, based on the Google data [31], we considered a reduction of (1-0.65)(1-0.2) (as compared with the period February 21-March 8) reflecting the draconian measures taken then. Based on the above, we attempted a forecasting of the pandemic in Lombardy from March 8 to May 4, the first day of the relaxation of the strict isolation measures.

## Results

As discussed, for our computations we ran 48 times the numerical optimization procedure for each combination of initial guesses based on the daily reported new cases from February 21 to March 8 and for each block of 48 runs, for further analysis we kept the values that yielded the smaller fitting error over all 48 runs. For the period February 21-March 8, which is used for the calibration of the model parameters, the median value of the ratio between the number of new cases of infected and recovered (excluding the zero values) is of the order of 10, while the average value of the ratio of the the number of new infected and deaths is of the order of 20. Hence, we have used as weights *w*_1_=1, *w*_2_=10, *w*_3_=20 to balance for the different scales of the number of infected vs. the number of recovered and dead. Other reasonable choices for the values of the weights around these values resulted in similar outcomes.

For all the near-optimal points obtained using the genetic algorithm optimization, the residuals were of the order of ~ 4,750,000. Regarding the values of the optimal parameters, we fitted their cumulative probability distributions using several functions, including the Normal, Log-normal, Weibull, Beta, Gamma, Burr, Exponential and Birnbaum-Saunders functions, and kept the one yielding the maximum Log-likelihood (see in the SI). Note that the optimal values of DAY-ZERO were between January 5 - January 23 (interquartile range, January 11 to January 18) (see Figure S1), the optimal values of *β* were between 0.636 and 0.86 (interquartile range, 0.665 to 0.719) (see Figure S2), and the optimal values of *∊* were between 0.01 and 0.249 (interquartile range, 0.023 to 0.1) (see Figure S3). The best fit to the distribution of optimal values of the DAY-ZERO was obtained using a Normal CDF with mean 1.67 (i.e. sim 2 days before the 16th of January) (95% CI:1.53, 1.80) and variance 4.43 (95% CI: 4.33, 4.52); thus taking the round value at 2 days, the expected DAY-ZERO corresponds to January 14 (interquartile range, January 11 to January 18).

The best fit to the distribution of the optimal values of *β*, was given by fitting a Burr CDF with *α* = 0.654 (95% CI: 0.653, 0.655), *c* = 276.061 (95% CI: 248.911, 306.172), *k* = 0.0537 (95% CI: 0.048, 0.060) having a mean value of 0.7 (interquartile range, 0.665 to 0.719). Finally, the best fit to the distribution of the optimal values of e was given by fitting a Birnbaum-Saunders CDF with parameters *μ* = 0.0492 (95% CI: 0.048, 0.0505 (scale parameter) and *α* = 0.934 (95% CI: 0.914, 0.954) (shape parameter), resulting in an expected value 0.07 (interquartile range, 0.023 to 0.1) (see in SI).

Thus, based on the derived values of the “effective” per-day disease transmission rate, the basic reproduction number *R*_0_ is 4.53 (min-max range: 4.40-4.65).

Finally, we ran the simulator for all values of the optimal triplets from the corresponding distributions as found by the solution of the optimization problem using the data from February 21 until March 8, then from March 9 to March 20 for validation purposes and from March 29 to May 4 for forecasting purposes.

Figures (2),(3),(4) depict the simulation results based on the optimal estimates, until the 19th of March. To validate the model with respect to the reported data of confirmed cases from March 9 to March 19, we have considered an (1-0.4)(1-0.2) reduction in the “effective” transmission rate and as initial conditions the values resulting from the simulation on March 8 as described in the methodology. Thus, the model approximated fairly well the dynamics of the pandemic in the period from February 21 to March 19.

**Figure 2:**
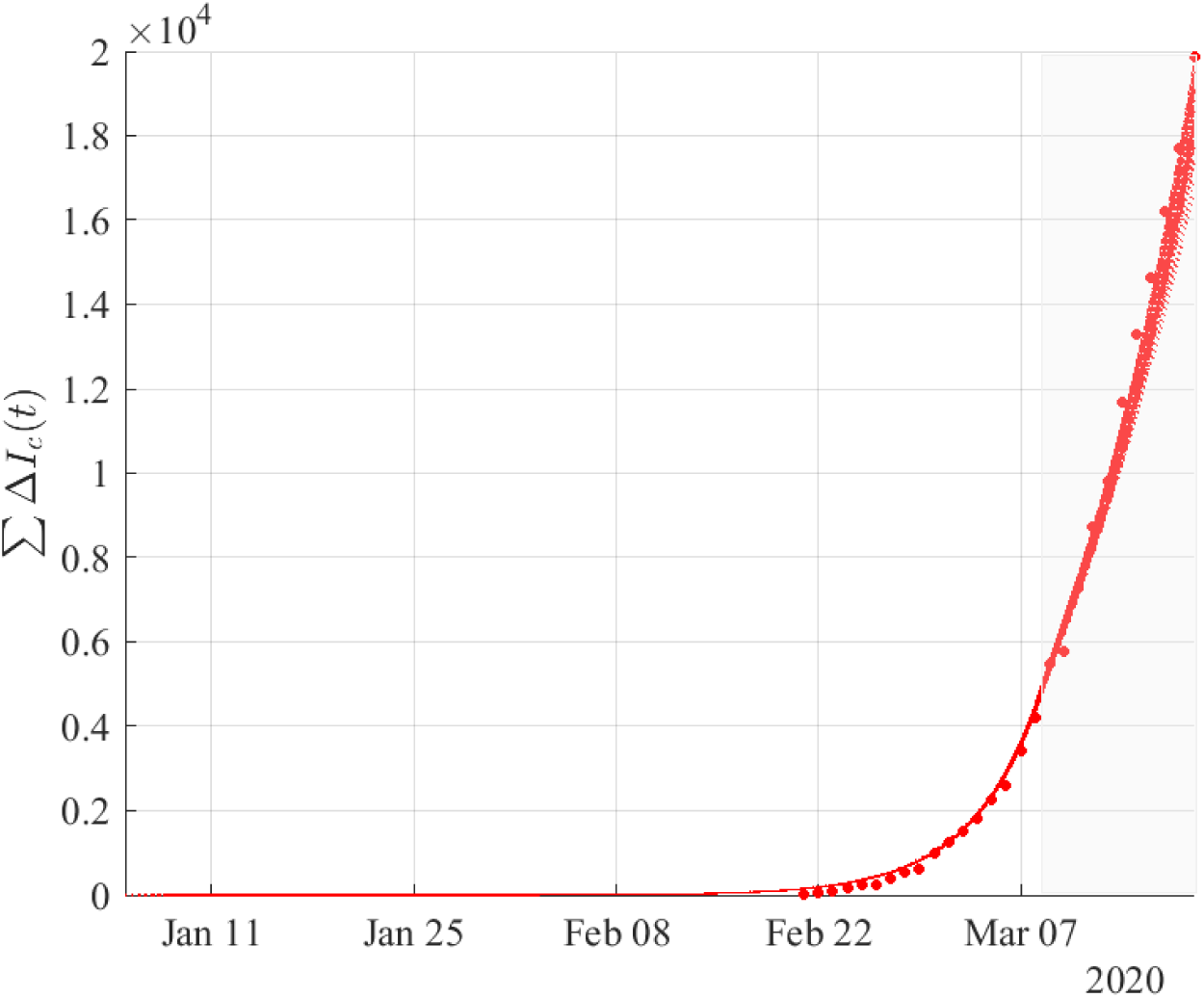
Cumulative number of infected cases from the compartment *I_c_* resulting from simulations based on the results obtained by fitting the daily new cases of infected from February 21 until the 8th of March. The validation of the model was performed using the reported data of confirmed cases from March 9 to March 19 (shaded area) by taking (1-0.4)(1-0.2) reduction in the “effective” transmission rate (see in Methodology) to the lockdown of March 8. Dots correspond to the reported data of confirmed cases.

**Figure 3:**
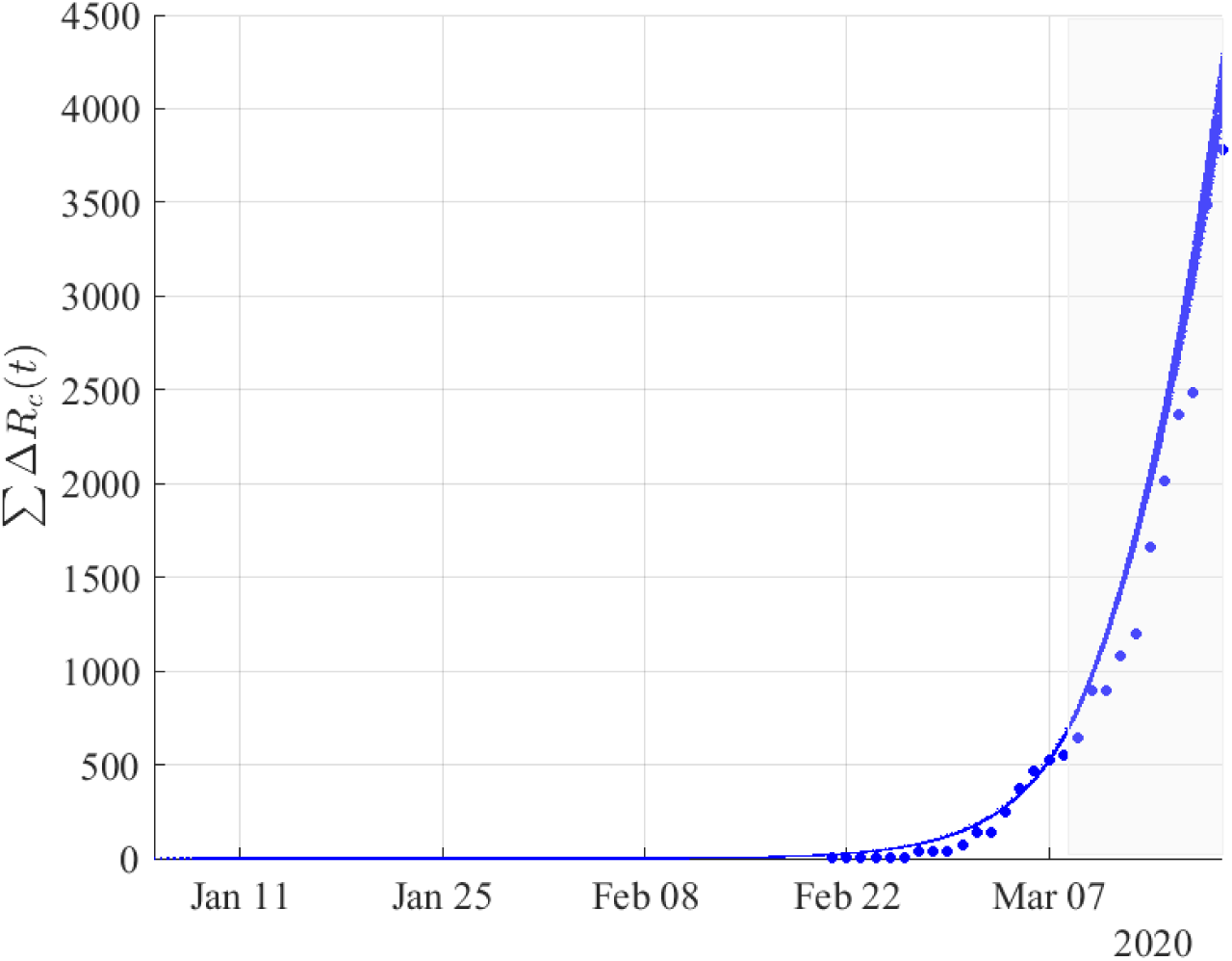
Cumulative number of recovered cases *R_c_*(*t*) resulting from simulations based on the results obtained by fitting the daily new cases of recovered from February 21 until the 8th of March. The validation of the model was performed using the reported data of confirmed cases from March 9 to March 19 (shaded area) by taking (1-0.4)(1-0.2) reduction in the “effective” transmission rate (see in Methodology) to the lockdown of March 8. Dots correspond to the reported data of confirmed cases of recovered.

**Figure 4:**
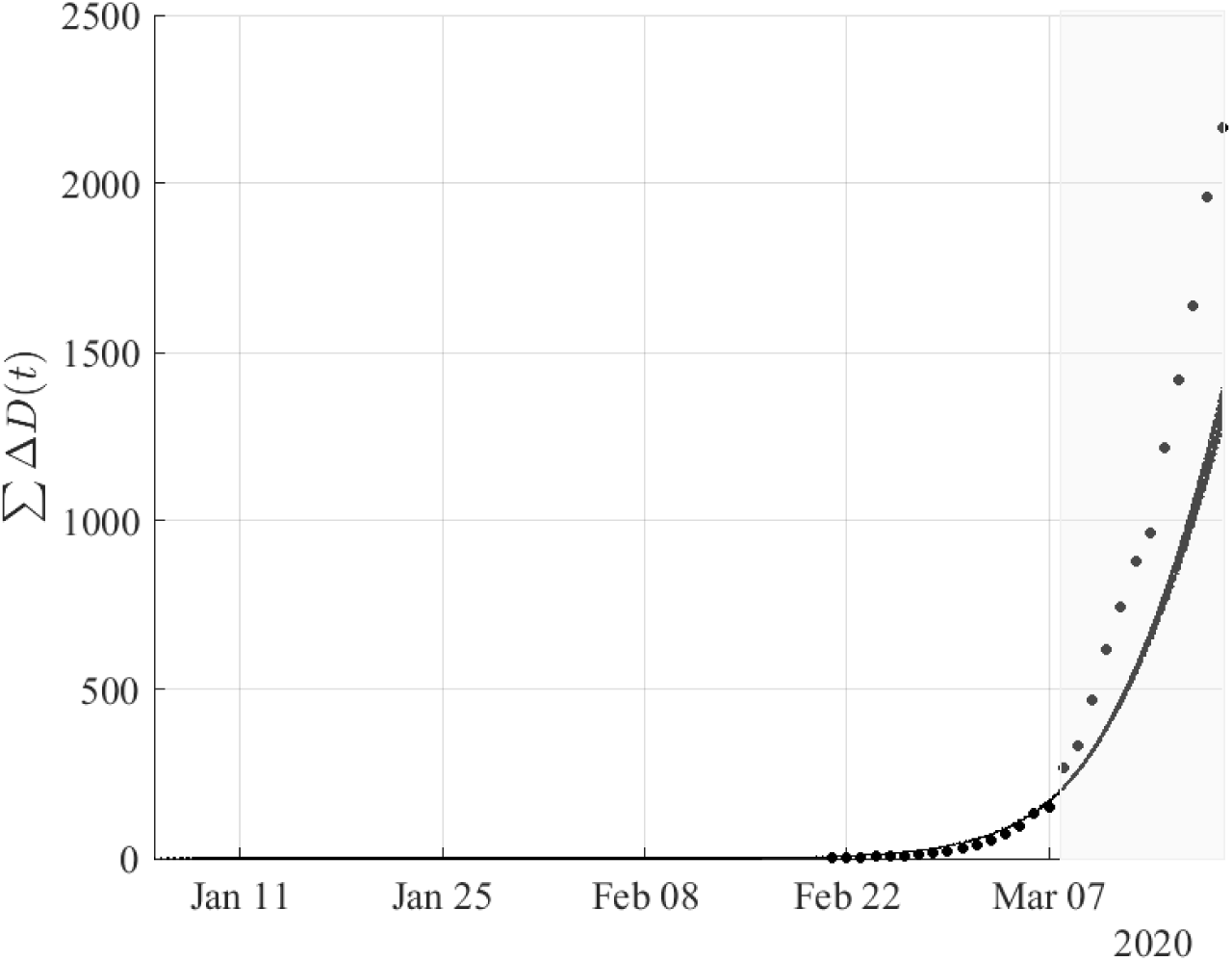
Cumulative number of deaths resulting from simulations based on the results obtained by fitting the daily new cases of deaths from February 21 until the 8th of March. The validation of the model was performed using the reported data of confirmed cases from March 9 to March 19 (shaded area) by taking (1-0.4)(1-0.2) reduction in the “effective” transmission rate (see in Methodology) to the lockdown of March 8. Dots correspond to the reported data of confirmed deaths.

As discussed in the Methodology, we also attempted based on our methodology and modelling approach to forecast the evolution of the outbreak until May 4, the first day of the relaxation of the measures. To do so, as described in the methodology, we have considered a (1-0.65)(1-0.2) reduction in the “effective” transmission rate starting on March 20 (compared to the period February 21-March 8), the day of announcement of even stricter measures in the region of Lombardy (see in Methodology). The result of our forecast is depicted in Figure 5. As shown, the model predicts quite well the evolution of the epidemic two months ahead of March 8 (the model parameters and the DAY-ZERO were estimated using the reported data from February 21 to March 8). Note that, regarding the confirmed cases, the mean values of *I_c_*(*t*) from over all simulations almost coincide with the reported values of the confirmed cases (see top panel of Figure 5). The reported recovered cases are in the lower part of the model predictions (*R_c_*(*t*)) (see medium panel of Figure 5). Also, the model predicts fairly the total number of deaths until May 4 (see bottom panel of Figure 5). A difference that can be observed with respect to the reported number of deaths and model forecasts in the period from March 18 to April 15 can be attributed to facts that the model did not take into account, such as the saturation of ICUs in that period, which could potentially lead to a larger number of deaths. Indeed, on March 8, 400 patients were in ICUs, on March 15, 700 patients (an increase of almost 80%), and after 6 days, on March 21, this number increased to 1093 corresponding to an increase of almost 150% with respect to the situation on March 8; the peak was on April 3, with 1381 people in ICUs. As a result, this period was the one with the highest numbers of reported deaths. Namely, the highest number of reported deaths in the region was on March 21 with 546 deaths, followed by ~540 deaths on March 27, ~40 deaths on March 28, ~415 deaths on March 29 and ~460 deaths on March 30. For comparison, on March 14 the reported number of deaths was 77, while the day after, on March 15, ~250 new deaths were reported, and on March 18, ~320 new deaths were reported. However, we note that the model was calibrated with the data reported until March 8. Until March 8, the case fatality ratio was of the order of ~9-10% and the number of cases admitted to ICUs was relatively small. After March 20 when there were many cases that were admitted to ICUs, the death toll increased significantly, it almost doubled, thus reaching the ~ 17-18% of the confirmed cases. Nevertheless, we must underline the very good agreement of the model predictions with the actual number of confirmed infected cases for the entire period March 8-May 4. This number, in contrast to the number of recovered and number of deaths, apparently is not shaped/biased by the capacity of ICUs.

**Figure 5:**
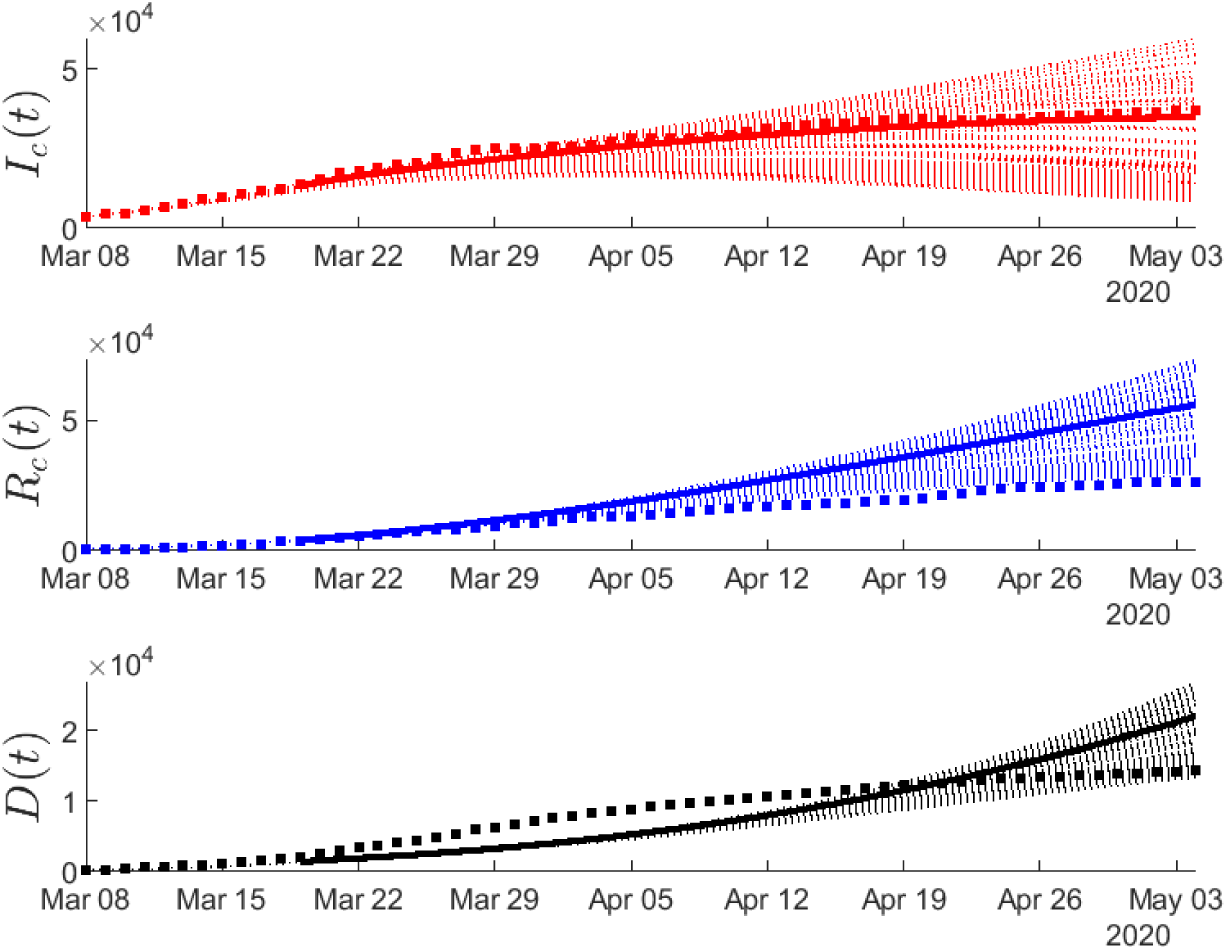
Estimated number of the infected (aka “confirmed” infected) cases with mild and severe symptoms (*I_c_*(*t*)), recovered (*R_c_*(*t*)) and deaths (*D*(*t*)) resulting from all simulations from March 8 (the day of the lockdown of all Italy) until May 4 (see in Methodology). Based on the Google released data (see in Methodology), we considered a (1-0.4)(1-0.2) reduction of the “effective” transmission rate from March 9 until March 19 compared to the period February 21-March 8, and then considering a (1-0.65)(1-0.2) reduction due to the even stricter mobility limitation measures announced by the government on March 20 and March 21 (see in Methodology). Red dots correspond to the reported confirmed infected cases, blue dots correspond to the reported recovered cases and black dots correspond to the reported deaths (up to May 4, 2020). Solid lines correspond to the mean values of *I_c_*(*t*), *R_c_*(*t*) and *D*(*t*), over all runs (see in Methodology).

## Discussion

The crucial questions about an outbreak is how, when (DAY-ZERO), why it started, and if and when it will end. Answers to these important questions would add critical knowledge to our arsenal to combat the pandemic. The tracing of DAY-ZERO, in particular, is of outmost importance. It is well known, that minor perturbations in the initial conditions of a complex system, such as the ones of an outbreak, may result in major changes in the observed dynamics. Undoubtedly, a high level of uncertainty for DAY-ZERO, as well as the uncertainty in the actual numbers of exposed people in the total population, raise several barriers to our ability to correctly assess the state and dynamics of the outbreak and to forecast its evolution and its end. Such pieces of information would lower the barriers and help public health authorities respond fast and efficiently to the emergency.

This study aimed exactly at shedding more light into this problem. To achieve this goal, we addressed a conceptually simple compartmental SEIIRD model with two infectious compartments in order to bridge the gap between the number of asymptomatic cases in the total population and the cases that will experience mild to more severe symptoms.

What is done until now with mathematical epidemiological models is the investigation of several scenarios, by changing e.g. the (or assuming a fixed) initial day (DAY-ZERO), the level of asymptomatic cases etc. Our work, is the first that introduces a methodological framework, to estimate the DAY- ZERO as well as the level of asymptomatic cases in the total population in a systematic way. Following the proposed methodological framework, we found that the DAY-ZERO in Lombardy was around the middle of January, a period that precedes by one month the fate of the first confirmed case in the hardest-hit northern Italian region of Lombardy. Interestingly enough, a recent study that was based on genomic and phylogenetic data analysis, reports the same time period, between the second half of January and early February, 2020, as the time when the novel coronavirus SARS-CoV-2 entered northern Italy [40]. Our analysis further revealed that the actual number of asymptomatic infected cases in the total population in the period until March 8 was around 15 times the number of confirmed infected cases, which until March 8 was also approximately equal to the number of cases that were hospitalized and admitted to ICUs.

Our model and methodological approach assume that there was one effective “ZERO” infected case that introduced the virus to the region; one could certainly argue that there were more than one cases that introduced the virus to the region on the same day; such scenarios can be investigated in a straightforward manner based on our proposed methodological approach. Furthermore, the proposed approach could be used for the quantification of the uncertainty of the evolving dynamics, taking into account the reported, from clinical studies, distributions of the epidemiological parameters rather than their expected values. We will consider this type of analysis in a future work.

Regarding the forecasting in Lombardy from March 8 until May 4 (the first day of relaxation of the measures), we have taken into account the very latest facts on the drop of human mobility, as released by Google [31] until April 30 for the region of Lombardy; these were shaped by the draconian measures announced on March 20-21 that included the closure of all parks, public and private offices and the prohibition of any pedestrian activity, even individually [39]. Our modelling approach approximated fairly well the dynamics of the pandemic in Lombardy two months ahead of time. The mean value of the evolution of the compartment that in our model reflects the confirmed cases, almost coincides with the reported cases for the entire period from March 8 to May 4.

To this end, we would like to make a final comment with respect to the basic reproduction number *R*_0_, the significance and meaning of which are very often misinterpreted and misused, thereby leading to erroneous conclusions. Here, we found a *R*_0_ ~ 4.5, which is higher compared to the values reported by many studies in China, and also in Italy, and in Lombardy in particular. For example, Zhao et al. estimated *R*_0_ to range between 2.24 (95% CI: 1.96, 2.55) and 3.58 (95% CI: 2.89, 4.39) in the early phase of the outbreak [8]. Similar estimates were obtained for *R*_0_ by Imai et al., 2.6 (95% CI: 1.5, 3.5) [5], Li et al. [41], Wu et al., 2.68 (95% CI: 2.47, 2.86), as well as by Anastassopoulou et al. recently, 3.1 (90% CI: 2.5, 3.7) [7].

Regarding Italy, D’Arienzo and Coniglio [42] used a SIR model to fit the reported data in nine Italian cities and found that *R*_0_ ranged from 2.43 to 3.10. In another study, the authors provided an estimate of the basic reproduction number by analyzing the first 5,830 laboratory-confirmed cases. By doing so they estimated the basic reproduction number at 3.1 [25].

First, we would like to stress that *R*_0_ is not a biological constant for a disease as it is affected not only by the pathogen, but also by many other factors, such as environmental conditions, demographics, as well as, importantly, by the social behavior of the population (see for example the discussion in [43]). Thus, a value for *R*_0_ that is found in one part of the world (e.g. in China), and even in a region of the same country, e.g. in Tuscany, Italy, cannot be generalized as a global biological constant for other parts of the world, or even for other regions of the same country. Obviously, the environmental factors and social behavior of the population in Lombardy are different from the ones, for example, prevailing in Hubei.

Second, most of the studies that provide estimates of the basic reproduction number are based solely on the reported cases, thus the actual number of infected cases in the total population, that may be asymptomatic but transmit the disease, is not considered; this fact may lead to an underestimation of the basic reproduction number. Moreover, in our approach as compared to clinical studies, the computation of *R*_0_ comes out as the necessary and sufficient condition of the stability as derived from the proposed model whose parameters are computed based on the available reported data, thus with a delay between the first actual case (see also the discussion in [44].

Our relatively simple conceptually model and approach do not aspire to accurately describe the complexity of the emergent dynamics, which in any case is an overwhelmingly difficult, if not impossible, task in the long run, even with the use of detailed agent-based models. We tried to keep the structure of the model as simple as possible in order to be able to model (in a coarse way) the uncertainty in both the “DAY-ZERO” and the number of asymptomatic actual cases in the total population using as few parameters as possible. The results of our analysis have indeed proved that the modelling approach succeeded in providing fair predictions of the evolution of the epidemic two months ahead of time. Such an early assessment would help authorities to evaluate the required measures to control the epidemic, such as the scale of diagnostic tests that have to be performed and the number of ICU required. While more complicated models can, in principle, be constructed to take into account more detailed information, such as the number of hospitalized patients and patients in ICUs, for any practical means such an approach would suffer from the “curse of dimensionality” as it would introduce many more parameters that would need calibration based on a relatively small size of data especially at the beginning of an outbreak. An attempt to compute the values of some of these additional parameters, which can only be roughly estimated by clinical studies at the early stages of an emerging novel infectious disease, would introduce additional uncertainty, thereby further complicating matters rather than solving the problem.

To this end, we hope that our conceptually simple, but pragmatic, modelling approach and methodological framework help to provide improved insights into the currently uncontrolled pandemic and to contribute to the mitigation of some of its severe consequences.

## Data Availability

The data used in this paper are given in the Supporting information.

## Author Contributions

Constantinos Siettos performed the formal numerical analysis and computations. Lucia Russo contributed to the development of the model and the formal analysis. Gennaro Nicola Bifulco analysed and collected the data. Gerardo Toraldo and Emilio Fortunato Campana contributed to the numerical analysis and optimization procedure. Cleo Anastassopoulou and Athanassios Tsakris interpreted the epidemiological meaning of the results. Constantinos Siettos, Cleo Anastassopolou and Lucia Russo wrote the manuscript. All authors reviewed the manuscript.

## Data Availability

All data referred to in the manuscript are provided as supplementary information.

## Supporting information

### Data

All the relevant data used in this paper are publicly available and accessible at http://www.salute.gov.it/. The reported cumulative numbers of cases from February 21 to March 19 are listed in Table S1. The reported numbers of confirmed cases from March 20 to May 20 are listed in Table S2.

**Table S1:** Reported cumulative numbers of cases for Lombardy, Italy (February 21-March 19)

| Calibration Data |  |  |  | Validation Data |  |  |  |
| --- | --- | --- | --- | --- | --- | --- | --- |
| Date | Infected | Recovered | Deaths | Date | Infected | Recovered | Deaths |
| Feb 21 | 15 | 0 | 0 | March 09 | 5469 | 646 | 333 |
| 22 | 54 | 0 | 1 | 10 | 5791 | 896 | 468 |
| 23 | 1101 | 0 | 6 | 11 | 7280 | 900 | 617 |
| 24 | 172 | 0 | 9 | 12 | 8725 | 1085 | 744 |
| 25 | 240 | 0 | 9 | 13 | 9820 | 1198 | 880 |
| 26 | 258 | 0 | 9 | 14 | 11685 | 1660 | 966 |
| 27 | 403 | 40 | 14 | 15 | 13272 | 2011 | 1218 |
| 28 | 531 | 40 | 17 | 16 | 14649 | 2368 | 1420 |
| 29 | 615 | 40 | 23 | 17 | 16220 | 2485 | 1640 |
| March 01 | 984 | 73 | 31 | 18 | 17713 | 3488 | 1959 |
| 02 | 1254 | 139 | 38 | 19 | 19884 | 3778 | 2168 |
| 03 | 1529 | 139 | 55 |  |  |  |  |
| 04 | 1820 | 250 | 73 |  |  |  |  |
| 05 | 2251 | 376 | 98 |  |  |  |  |
| 06 | 2612 | 469 | 135 |  |  |  |  |
| 07 | 3420 | 524 | 154 |  |  |  |  |
| 08 | 4189 | 550 | 267 |  |  |  |  |

**Table S2:** Reported numbers of confirmed cases for Lombardy, Italy (March 20-May 4)

| Date | Infected | Recovered | Deaths | Date | Infected | Recovered | Deaths |
| --- | --- | --- | --- | --- | --- | --- | --- |
| March 20 | 15420 | 4295 | 2549 | April 16 | 33090 | 18396 | 11608 |
| 21 | 17370 | 5050 | 3095 | 17 | 33434 | 18850 | 11851 |
| 22 | 17885 | 5865 | 3456 | 18 | 34195 | 19136 | 12050 |
| 23 | 18910 | 6075 | 3776 | 19 | 34497 | 19526 | 12213 |
| 24 | 19868 | 6657 | 4178 | 20 | 34587 | 20008 | 12376 |
| 25 | 20591 | 7281 | 4474 | 21 | 33978 | 21374 | 12579 |
| 26 | 22189 | 7839 | 4861 | 22 | 34242 | 22110 | 12740 |
| 27 | 23895 | 8001 | 5402 | 23 | 33873 | 23352 | 12940 |
| 28 | 24509 | 8962 | 5944 | 24 | 34368 | 23782 | 13106 |
| 29 | 25392 | 9255 | 6360 | 25 | 34473 | 24227 | 13269 |
| 30 | 25006 | 10337 | 6818 | 26 | 35166 | 24398 | 13325 |
| 31 | 25124 | 10885 | 7199 | 27 | 35441 | 24589 | 13449 |
| April 1 | 25765 | 11415 | 7593 | 28 | 35744 | 25029 | 13575 |
| 02 | 25876 | 12229 | 7960 | 29 | 36122 | 25333 | 13679 |
| 03 | 26189 | 13020 | 8311 | 30 | 36211 | 25749 | 13772 |
| 04 | 27220 | 13242 | 8656 | May 1 | 36473 | 26136 | 13860 |
| 05 | 28124 | 13426 | 8905 | 2 | 36667 | 26146 | 14189 |
| 06 | 28469 | 13863 | 9202 | 3 | 36926 | 26371 | 14231 |
| 07 | 28343 | 14498 | 9484 | 4 | 37307 | 26504 | 14294 |
| 08 | 28545 | 15147 | 9722 |  |  |  |  |
| 09 | 29074 | 15706 | 10022 |  |  |  |  |
| 10 | 29530 | 16280 | 10238 |  |  |  |  |
| 11 | 30258 | 16823 | 10511 |  |  |  |  |
| 12 | 31265 | 17166 | 10621 |  |  |  |  |
| 13 | 31935 | 17478 | 10901 |  |  |  |  |
| 14 | 32363 | 17821 | 11142 |  |  |  |  |
| 15 | 32921 | 17855 | 11377 |  |  |  |  |

### Fitting the Distributions of the Optimal Values

We fitted the cumulative probability distributions of DAY-ZERO *β*, *∊* using several functions including the Normal, Log-normal, Weibull, Beta, Gamma, Burr, Exponential and Birnbaum-Saunders CDFs and kept the one resulting in the maximum Log-likelihood. For the DAY-ZERO, the best fit to the distribution of the optimal values was obtained by the Normal CDF:

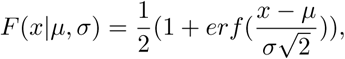

with mean value *μ* = 1.67 (95% CI: 1.53, 1.80) and *σ* = 4.43 (95% CI:4.33, 4.52).

The cumulative distribution of DAY-ZERO and the resulting exponential distribution are given in Figure S1.

For the distribution of *β* values the best fitting distribution was obtained by the Burr CDF which is a three-parameter family of curves given by:

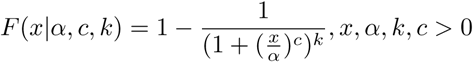

with *α* = 0.654 (95% CI: 0.653, 0.655), *c* = 276.061 (95% CI: 248.911, 306.172), *k* = 0.0537 (95% CI: 0.048, 0.060). Thus, the resulting mean value is *β* = 0.70. The cumulative distribution of *beta* and the resulting Burr CDF distribution are given in Figure S2.

For *∊*, the best fit to the distribution of the optimal values was obtained by a Birnbaum-Saunders CDF:

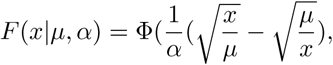

with *μ* = 0.0492 (95% CI: 0.048, 0.0505) (scale parameter) and *α* = 0.934 (95% CI: 0.914, 0.954) (shape parameter). Φ(*x*) denotes the standard normal CDF. The mean value is given by:

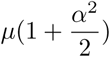

**Figure S1:**
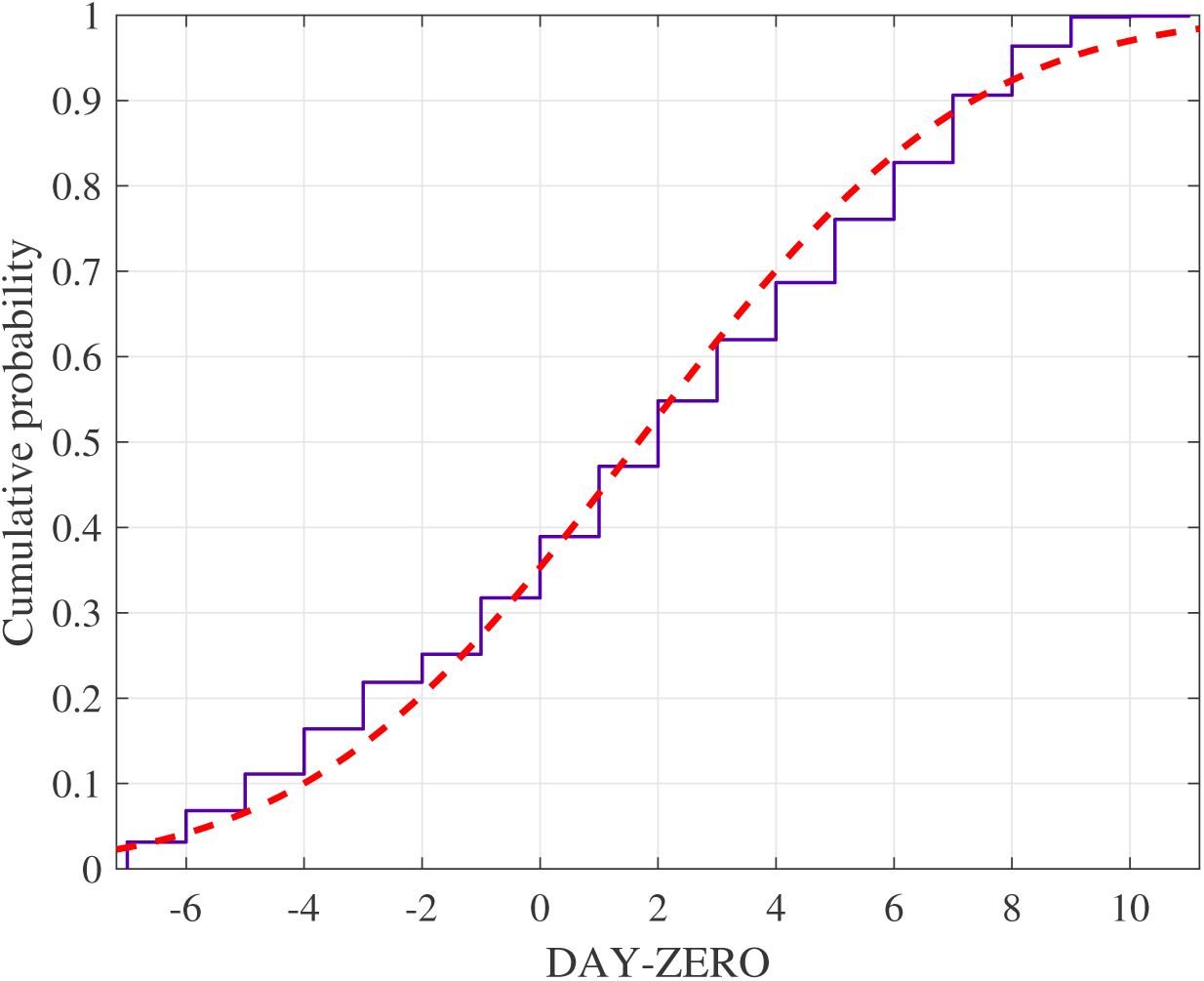
Cumulative probability distribution of the optimal values of DAY-ZERO as obtained by running the optimization problem using a grid of 20 × 13 × 15 initial guesses, thus using a 2 days step for the DAY-ZERO within the interval 27 December 2019-5th of February 2020 i.e. ±20 days around the 16th of January, a step of 0.05 within the interval (0.3, 0.9) for and a step of 0.02 within the interval (0.01, 0.29) for *∊* =. The best fit was obtained with the Normal CDF with *μ* = 1.67 (95% CI: 1.53, 1.80) (thus corresponding to the 14th of January) and *σ* = 4.43 (95% CI:4.33, 4.52).

**Figure S2:**
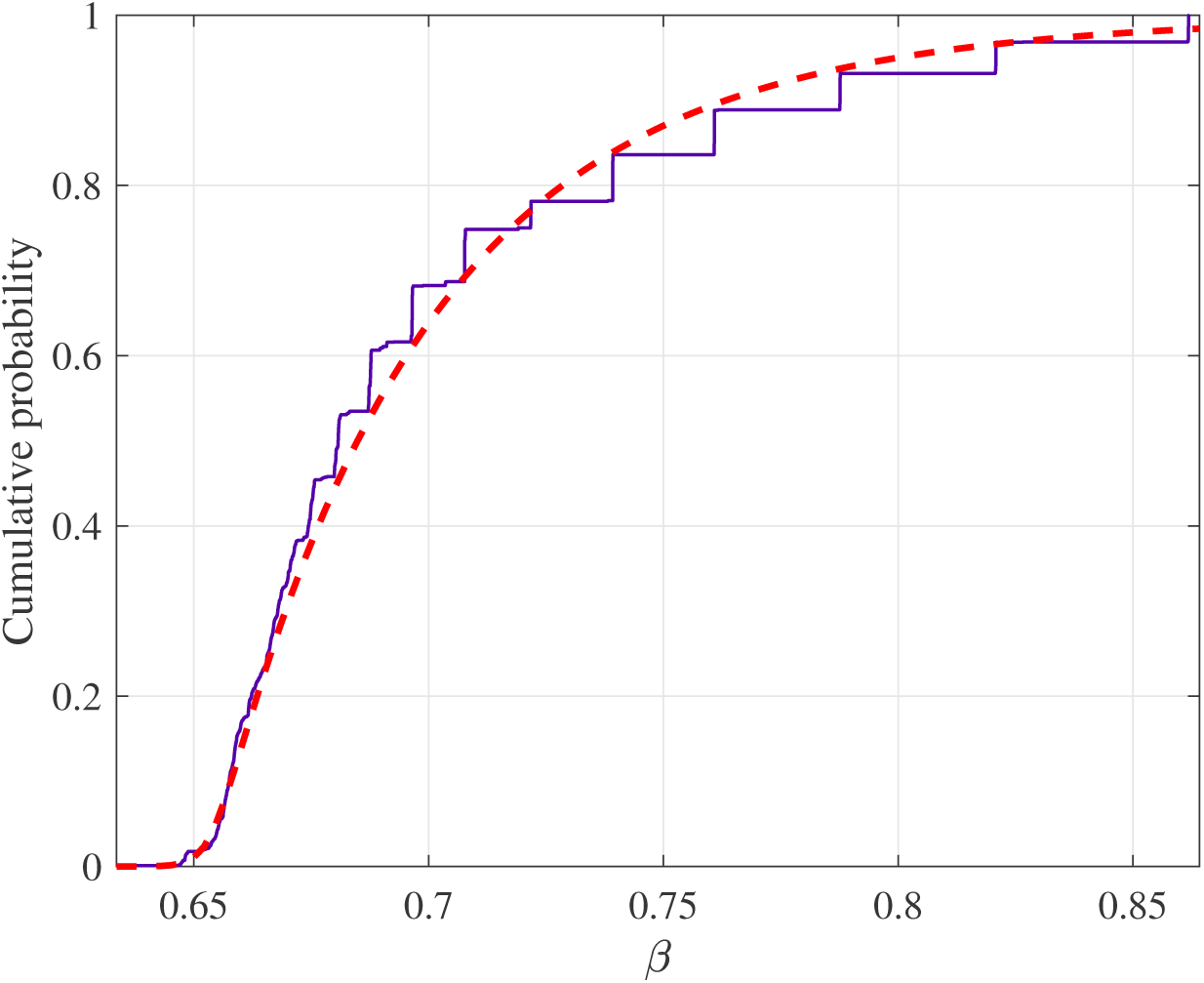
Cumulative probability distribution of the optimal values of *β* as obtained by running the optimization problem using a grid of 20 × 13 × 15 initial guesses, thus using a 2days step for the DAY-ZERO within the interval 27 December 2019-5th of February 2020 i.e. ±20 days around the 16th of January, a step of 0.05 within the interval (0.3, 0.9) for *β* and a step of 0.02 within the interval (0.01, 0.29) for *∊*. The best fit was obtained with the Burr CDF, with *α* = 0.654 (95% CI: 0.653, 0.655), *c* = 276.061 (95% CI: 248.911, 306.172), *k* = 0.0537 (95% CI: 0.048, 0.060).

**Figure S3:**
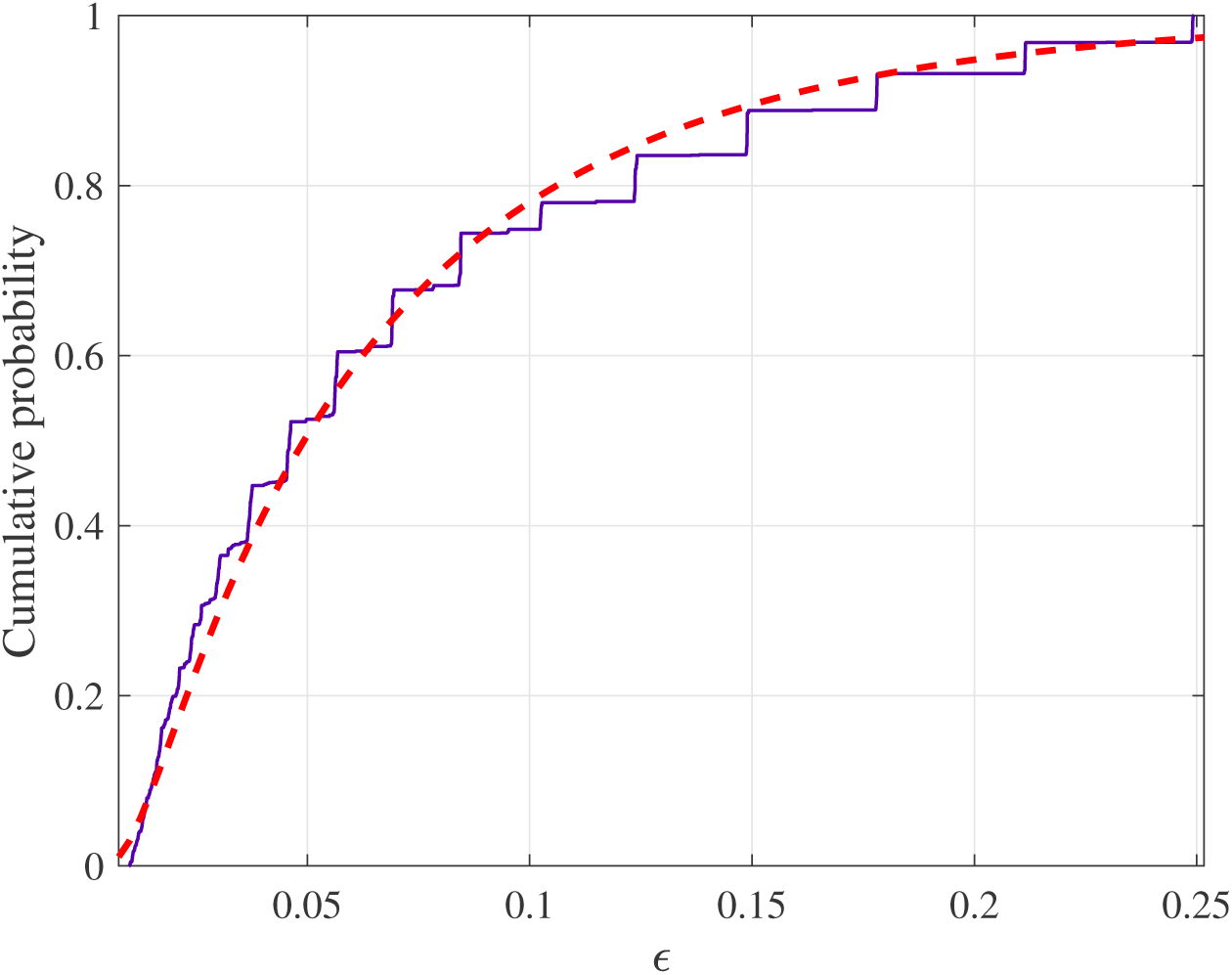
Cumulative probability distribution of the optimal values of *∊* as obtained by running the optimization problem using a grid of 20 × 13 × 15 initial guesses, thus using a 2days step for the DAY-ZERO within the interval 27 December 2019-5th of February 2020 i.e. ±20 days around the 16th of January, a step of 0.05 within the interval (0.3, 0.9) for *β* and a step of 0.02 within the interval (0.01, 0.29) for *∊*. The best fit was obtained by a Birnbaum-Saunders CDF, *μ* = 0.0492 (95% CI: 0.048, 0.0505) (scale parameter) and *α* = 0.934 (95% CI: 0.914, 0.954) (shape parameter).

Thus, the mean value is given by *∊* = 0.0707.

The cumulative distribution of ɛ and the resulting exponential distribution are given in Figure S3.

